# Predicted impact of the COVID-19 pandemic on global tuberculosis deaths in 2020

**DOI:** 10.1101/2020.04.28.20079582

**Authors:** Philippe Glaziou

## Abstract

Policies widely adopted in response to the ongoing pandemic of Covid-19, particularly lockdowns and reassignments of health personnel and equipment, are impacting the performance of TB prevention and care programmes. Estimates of the impact of reductions in the performance of global TB detection and care on TB mortality over 2020 are presented. A global TB case detection decrease by an average 25% over a period of 3 months (as compared to the level of detection before the pandemic), will lead to a predicted additional 190 000 (56 000 – 406 000) TB deaths.

---

Policies widely adopted in response to the ongoing pandemic of Covid-19^1^, particularly lockdowns and reassignments of health personnel and equipment, are impacting the performance of TB prevention and care programmes at a level varying significantly between countries.

In China, national TB case detection in February 2020 dropped by 20% in comparison with the number of cases detected in February 2019. In India (Figure 1), weekly counts of reported cases dropped by 75% in the three weeks following 22 March (average 11367 weekly cases), the date of a strict national lockdown implementation, compared to an average of 45875 weekly cases during the previous weeks of 2020, a drop attributable to a combination of factors including delays in entering the data onto the real-time national online TB surveillance system Nikshay^2^, reduced attendance to health services, reassignment of health personal and a reduction in TB testing and detection. Similarly, case reporting dropped recently by 68% in January-March in Indonesia. In contrast, the national TB programme in Brazil reported no recent change in weekly case counts at the national level. A small number of TB patients were reported with confirmed Covid-19 disease, but at the date of this report, the extent of the size of the dual epidemic is largely unknown.

**Figure 1.**
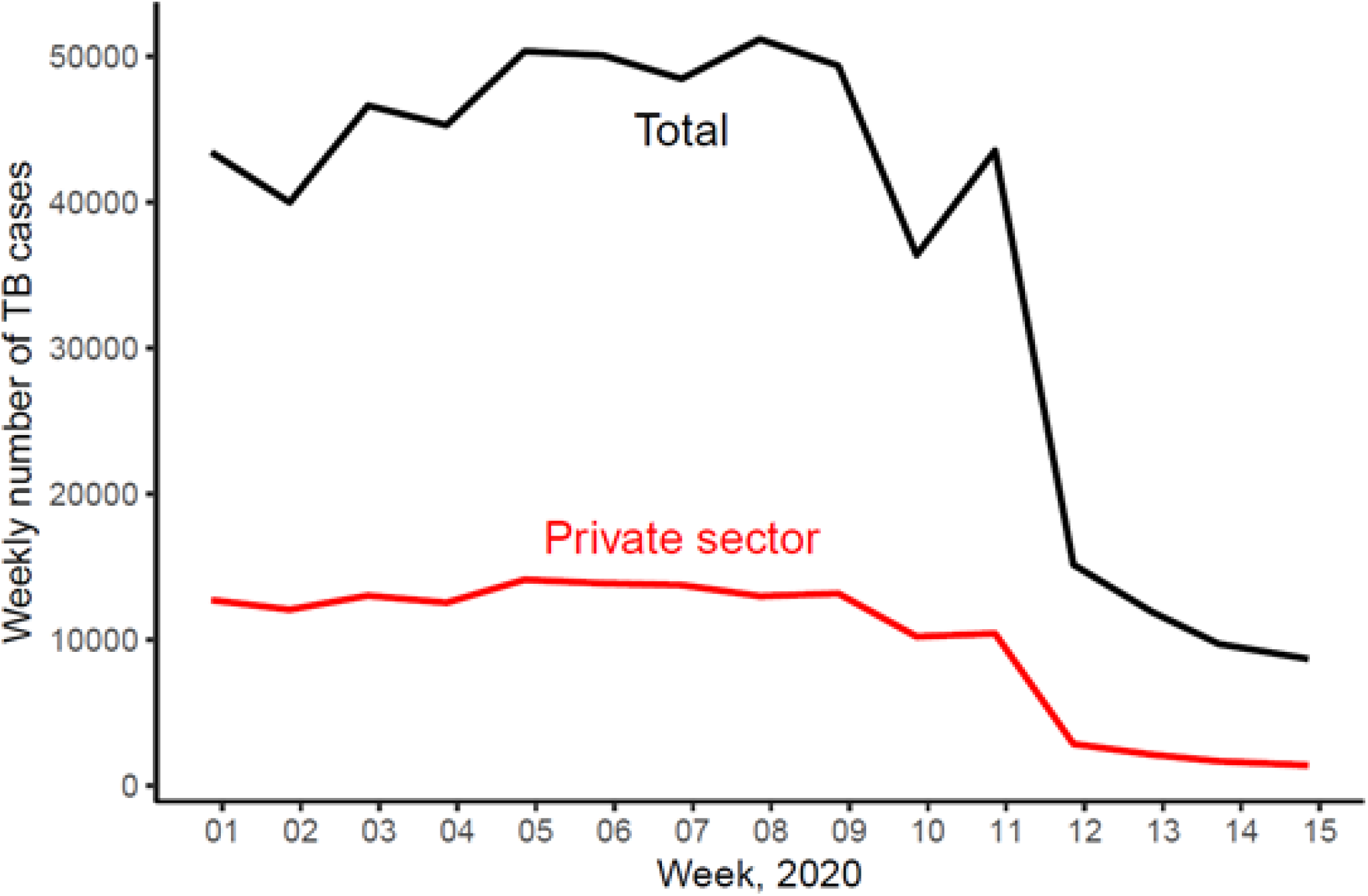
Trends in weekly case notifications in India in 2020

Based on the above observations and similar country reports, it can be predicted that if TB detection and care services are not maintained at the level of performance before the Covid-19 pandemic started, TB case detection will temporarily drop at the global level, with a resulting increase in TB mortality primarily affecting the most vulnerable TB patients.

The impact of reductions in the performance of TB detection and care on TB mortality over 2020 is modelled using methods described in appendix, under varying levels of decrease in global case detection over a varying duration. The model assumes that TB incidence in 2020 continues its slow decline from previous years and its trends are not significantly affected in the short term by the Covid-19 pandemic. The beneficial effect of lockdown and social distancing policies on the TB reproductive number may be offset by increased duration of infectiousness under lower case detection and treatment performance. In the absence of clinical data, no assumption is made about increased average case fatality rates among dually infected Covid-TB individuals (with likely conservative estimates of mortality) and deaths among Covid-TB coinfected individuals are attributed to TB as the underlying cause.

Figure 2 shows the predicted number of additional TB deaths (including TB deaths in HIV-negative and in HIV-positive individuals) that will result from a temporary decrease in global TB case detection. The estimated additional TB deaths are in excess of an expected 1.47 (uncertainty interval 1.1 – 1.9) million TB deaths that would have been predicted in the absence of the Covid-19 pandemic, based on recent trends prior to 2020. For example, if global TB case detection decreases by an average 25% over a period of 3 months (as compared to the level of detection before the pandemic), an additional 190 000 (56 000 – 406 000) TB deaths are predicted.

**Figure 2.**
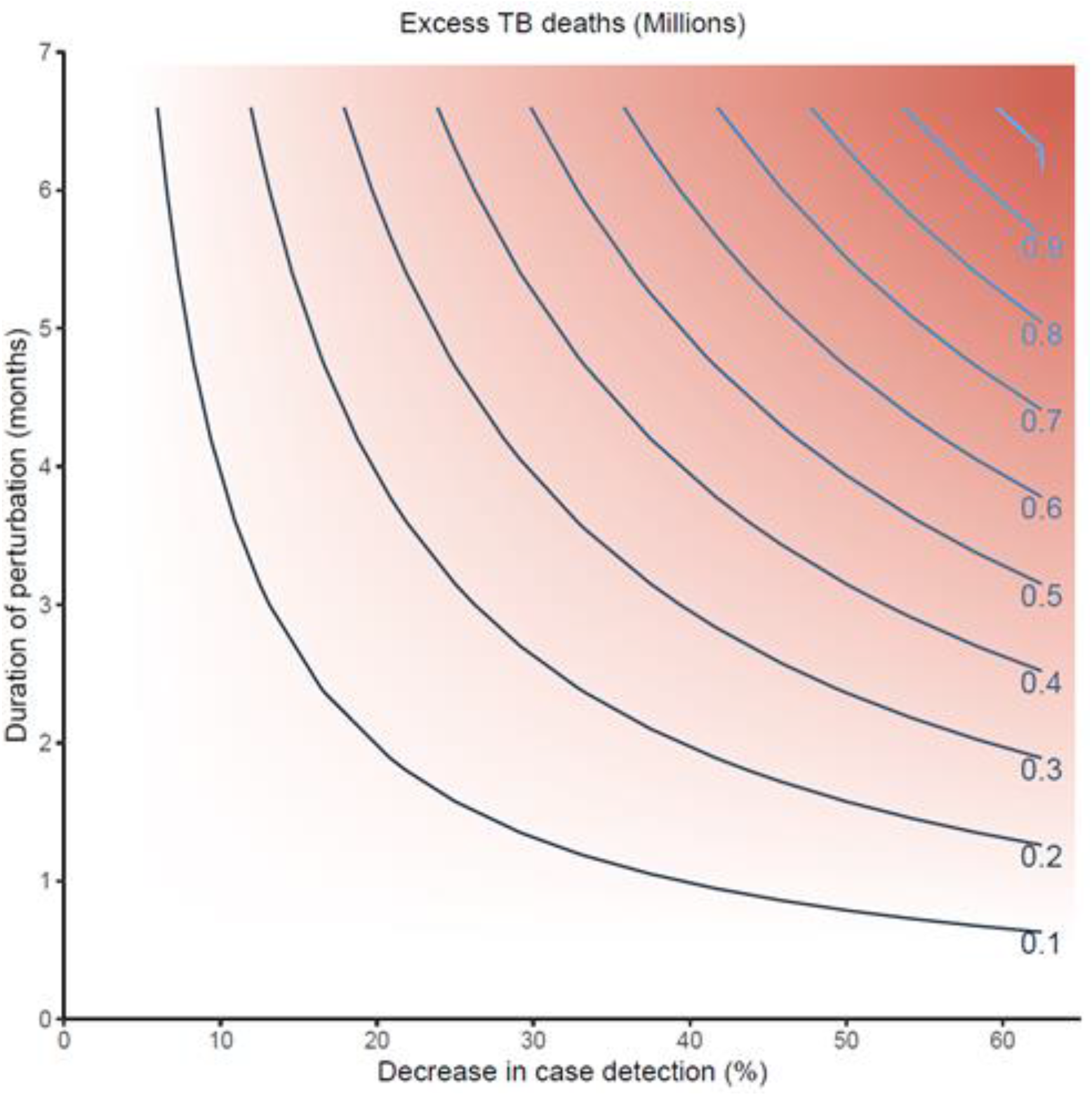
Predicted excess in TB deaths in 2020 in relation to the duration and extent of a temporary average reduction in TB case detection

The predicted impact on TB mortality in 2020 highlights the urgency to monitor weekly counts of newly reported TB cases in countries with real-time TB case reporting, and monthly counts in other countries, and to promptly restore the functionality of negatively affected TB detection and care programmes. National TB detection and care programmes should be considered essential health services to prioritize and maintain during the covid-19 pandemic.

Further modelling is required for more detailed and longer-term predictions of the impact of the Covid-19 pandemic on the burden of TB and its determinants, particularly undernutrition, and on progress towards targets set at the UN General Assembly High-Level Meeting on TB in September 2018. The response to the Covid pandemic may bring opportunities for synergies including increased levels of TB testing, particularly in high-HIV settings where symptoms of TB and Covid-19 disease are more difficult to differentiate clinically, better implementation of infection control measures and more effective contact tracing investigations.

## Data Availability

All model input data are publicly available.

https://www.who.int/tb/country/data/download/en/

## Appendix

The perturbation *d* of TB detection and care is expressed in terms of a reduction in the number of treated cases as compared to the expected number in the absence of the Covid-19 pandemic.

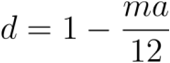

where *a* is the average monthly drop in case detection (0 ≤ *a* ≤1) compared to the expected value in the absence of a perturbation and *m* the number of months during which case detection is affected in 2020 (0 ≤ *m* ≤ 12). The perturbation is null at *d* = 1 and maximum at *d* = 0.

TB mortality *M*_*d*_ is obtained as follows

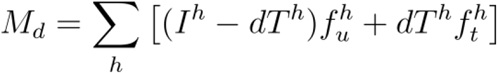

where *I* denotes incident TB cases, *T* the number of treated cases, *f* the case fatality rate, superscript *h* denotes HIV status (infected or not), subscript *u* denotes untreated TB and subscript *t* denotes treated TB. Distributions of *f* are taken from the technical appendix of Global TB Report 2019^3^. It is assumed that the value for *d* is the same among HIV-positive and HIV-negative individuals. Estimates are conservative if the value of *d* is lower among HIV-positive individuals, however, most of the global TB mortality is among HIV-negative individuals.

In the absence of a rebound in case detection above values prior to the perturbation, the excess in TB mortality Δ*M* resulting from the perturbation *d* is thus

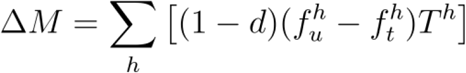

Errors in real-valued random variates *I* and *f* are propagated by approximating a function *g*(.) using second-order Taylor series expansion about its moments.

